# Theory of Mind Variability in Schizophrenia: A Neurodevelopmental Perspective through Neurological Soft Signs and Premorbid Adjustment

**DOI:** 10.1101/2024.12.18.24319277

**Authors:** M. Giralt-López, S. Miret, S. Campanera, M. Moreira, A. Sotero-Moreno, N. Hostalet, L. Lázaro, MO. Krebs, L. Fañanás, M. Fatjó-Vilas

## Abstract

**Background:** Theory of Mind (ToM) is impaired in individuals with schizophrenia (SZ). Given the neurodevelopmental nature of both social cognition and SZ, variations in ToM abilities likely originate early in life. Thus, indirect markers of altered neurodevelopment, such as neurological soft signs (NSS) and premorbid adjustment (PA), may help explain these differences.

**Methods:** The study included 38 patients with schizophrenia-spectrum disorder (SSD), 26 healthy siblings and 47 controls. ToM was assessed using the Hinting Task (HT). NSS were evaluated with the Neurological Evaluation Scale (NES) and PA with the Premorbid Adjustment Scale (PAS), yielding Social and Academic scores. Intelligence Quotient (IQ) was estimated two subtests of the WAIS-III and Family History (FH) through the Family Interview for Genetic Studies (FIGS).

**Results:** First, patients presented more deficits in two subscales of the NES (motor coordination and sequencing of complex motor acts) than siblings and controls, with siblings performing intermediate in the sequencing subscale (linear mixed models, adjusted for age, sex and IQ). Patients showed worse social PA than siblings during childhood and late adolescence. Second, patients showed poorer HT performance than siblings and controls, but the neurodevelopmental markers did not modulate such differences. Third, within each group, neurodevelopmental vulnerability markers were not associated with ToM performance (linear regression analyses adjusted for age, sex, IQ and FH).

**Conclusion:** In our sample, while patients showed more evidence of neurodevelopmental deviances than siblings and controls, such differences did not contribute to ToM variability, suggesting that they may stem from different neurodevelopment-related pathways.

## 1. INTRODUCTION

Social cognition refers to a wide range of skills that allow people to perceive, interpret and process social stimuli, guiding social interactions. It is a multi-dimensional construct that comprises functions such as emotional processing, social perception and knowledge, theory of mind and attributional bias (Green et al., 2008). Among all these social cognition dimensions, the current article focuses on the theory of mind (ToM, also known as "mentalising"). It is defined as the ability to deduce the mental states of others, such as their beliefs, intentions, desires, and emotions (Frith & Frith, 1999).

The development of ToM skills includes, first, the mental construction of the object as existing outside and independent of the subject, the recognition of himself as separated from the others and the engagement in joint attention activities with others or the pretence play. Next, between 3 and 4 years of age, the child can infer that another person may have a belief different from its own, called "first-order ToM" (Baron-Cohen et al., 1985). Later, between 6-and 7 years old, the child begins to understand that the other person can also represent other people’s mental state, known as "second-order ToM" (Damasceno, 2020). As ToM performance continues to improve throughout childhood and adolescence into adulthood (Meinhardt-Injac et al., 2020), proper brain maturation likely plays a crucial role in acquiring and performing these skills.

Although the precise cerebral organisation of social processes is not fully defined, specific brain regions and networks are associated with social information processing (Gazzaniga MS, 2019; Green et al., 2015; Magno et al., 2022). In particular, the medial prefrontal cortex (PFC) is involved in interpreting nonverbal social cues, understanding complex social contexts, inferring others’ internal states and beliefs, and evaluating their traits over time, skills closely related to ToM performance (Kozhuharova et al., 2020). From an evolutionary standpoint, the PFC’s involvement in social cognitive abilities such as ToM aligns with the idea that the expansion of the cerebral cortex is the substrate for human-specific higher cognitive processes (Sousa et al., 2017). It is important to note that while our closest primate relatives exhibit essential components of ToM (i.e., understanding goals, intentions, and perceptions), the ability to understand that others can hold beliefs that are incorrect or misaligned with reality appears exclusively human. (Call & Tomasello, 2008).

From a developmental view, the gradual achievement of ToM skills from infancy through childhood into adulthood aligns with the maturation of the PFC `(Blakemore& Choudhury, 2006)`. For this reason, the PFC is viewed as a region particularly susceptible to disruption due to its long-lasting period of maturation (in humans, not complete until near the age of 25). Additionally, researchers hypothesize that brain regions, such as the PFC, which evolved most recently in humans, play a crucial role in advanced cognitive abilities, such as high-order ToM (i.e., the ability to infer hidden intentions through indirect speech or to grasp false beliefs). Interestingly, these same regions are involved in different neurodevelopmental disorders, such as schizophrenia and autism, which involve impairments in ToM (Teffer & Semendeferi, 2012).

Accordingly, ToM impairments are well-documented in schizophrenia (SZ) (Bora & Pantelis, 2013; Brune, 2005; Chung et al., 2014; Giralt-López et al., 2020; Mondragón-Maya et al., 2017). Some studies interpret these deficits as a state marker linked to symptomatology (Balogh et al., 2014; Mazza et al., 2012). Others, however, describe ToM impairments as a trait marker, noting that they are not influenced by clinical severity or insight (Ay et al., 2016; Giralt-López et al., 2020; Sprong et al., 2007) and are stable over time, as seen in prospective data (Ayesa-Arriola et al., 2014). Supporting the trait marker view, ToM deficits have also been observed in first-episode psychosis (FEP) and individuals at clinical or genetic high risk for psychosis (unmedicated prodromal subjects or first-degree relatives, respectively) (Bora & Pantelis, 2013; Healey et al., 2016). Based on these data and considering the developmental nature of ToM, it is appropriate to examine the variability in ToM impairments in patients and family members through the lens of the neurodevelopmental model of SZ. This model suggests that SZ arises from abnormal neurodevelopmental processes, starting years before the illness manifests, due to the complex interaction of genetic susceptibility and environmental factors (Birnbaum & Weinberger, 2017; Jones & Murray, 1991; Rapoport et al., 2012).

Clinical data have also shown that children who later develop SZ face earlier developmental, educational, and social challenges compared to others (Jaaro-Peled & Sawa, 2020) exhibit neuromotor dysfunction (Cunningham Owens & Johnstone, 2006; Rosso et al., 2000; Walker et al., 1994), and worse general cognitive functioning (Cannon et al., 2006; Sheffield et al., 2018) all contributing to poorer premorbid adjustment (PA) (Parellada et al., 2017). In turn, poor PA was significantly associated with prominence of negative symptoms and worse quality of life functional outcome (MacBeth & Gumley, 2008), early age of onset, educational problems, chronicity, and neurological soft signs (NSS), becoming an essential predictor of a particularly severe form of SZ (Gupta et al., 1995).

In this sense, NSS are early indirect markers of subtle deviations from normal neurodevelopment that have been associated with SZ (Bora et al., 2018; Tsapakis et al., 2023). They are minor neurologic deficits observable by clinical examination, including deficits in sensory integration, motor coordination, sequencing of complex motor acts, eye movements, and developmental reflexes. NSS occur more frequently and pronounced in SZ than in other neuropsychiatric disorders (Bora et al., 2018), with a prevalence of over 50% compared to about 5% in healthy individuals (Rathod et al., 2020a). Also, NSS are more common in childhood and adolescent-onset SZ than in adult-onset cases (Biswas et al., 2007) and correlate with negative symptoms and cognitive dysfunction in SZ patients (Chan et al., 2015). This suggests a relationship between NSS and deficit schizophrenia in a subgroup of patients with neurodevelopmental alterations (Hoffmann et al., 2018).

NSS are present in prodromal phases before medication exposure (Kong et al., 2019), first-episode and chronic SZ (Chan, Xu, Heinrichs, Yu, & Wang, 2010a), and are more prevalent in first-degree relatives of people with SZ than in controls (Chan, Xu, Heinrichs, Yu, & Gong, 2010; Feng et al., 2020; Neelam et al., 2011). Furthermore, NSS were found to have moderate but significant heritability in a healthy twin sample and patients with SZ showed a strong correlation in NSS with their first-degree relatives (Xu et al., 2016), which supports a trait perspective and their potential endophenotypic role (Chan & Gottesman, 2008). However, evidence shows that NSS fluctuate, decreasing as psychopathological symptoms remit, though they do not return to levels seen in healthy controls, generating a state-trait debate about NSS (Bachmann & Schröder, 2018).

Considering all the points mentioned above, it can be hypothesised that the variability in ToM abilities observed after the onset of the illness likely originates early in life. Therefore, the presence or absence of indirect markers of altered neurodevelopment, such as NSS or distinct patterns of PA, might play a key role in shaping these differences. In other words, these early indicators could reflect distinct neurodevelopmental trajectories that influence how ToM abilities evolve, potentially affecting how the illness manifests and progresses in different individuals.

Some previous studies have explored the relationship between NSS and PA and ToM, but they are scarce, and none explored this relation in non-affected relatives (Herold et al., 2019; Punsoda-Puche et al., 2024; Romeo et al., 2014). Then, we aimed to deepen our understanding of ToM variability and its potential role as a trait marker by examining whether ToM performance could be better characterised by considering other markers of altered neurodevelopment, such as poorer PA and the severity of NSS in patients, siblings and controls.

## 2. METHODS

### 2.1 Sample

The sample included 121 participants: 38 patients diagnosed with recent onset schizophrenia-spectrum disorder (SSD), 26 healthy siblings from these patients and 47 controls. All participants were of European origin. The sample was recruited at the Outpatient Mental Health Clinic of the Hospital Santa Maria de Lleida and assessed when clinically stable by the same clinician (SM). It is important to note that the sample in this study partially overlaps with a dataset previously analysed by Giralt-López et al. (2020 and 2024). In this research, we integrated methodologies from our prior studies. First, we have focused specifically on siblings as the relatives’ group, to reduce the heterogeneity of this group and some confounding factors (such as age or others related to the cohort effect). Second, we have included a control sample, which was recruited in Lleida and Barcelona from non-medical staff working in the hospital, their relatives and acquaintances, and independent community sources.

The key contribution and originality of this study lie in combining neurodevelopmental markers to better understand ToM variability.

Patients were diagnosed according to DSM-IV-TR criteria and interviewed using the Comprehensive Assessment of Symptoms and History (CASH) (Andreasen et al., 1992). Siblings were evaluated through a clinical interview and the Structured Clinical Interview for DSM Disorders (SCID) (First & Gibbon, 2004), and only those who had no history of current or lifetime psychotic spectrum disorders or mood disorders were included in the study. The control group did not have a personal or family history of psychiatric disorders or any prior treatments for such conditions. Other common exclusion criteria for all groups were intellectual disability, any severe medical conditions that could impair brain function, neurological disorders, or previous head injuries with loss of consciousness.

### 2.2 Assessments

Social cognition, specifically ToM, was assessed through the Spanish version of the Hinting Task (HT) (Corcoran et al., 1995; Gil et al., 2012), a test comprising ten brief stories involving two people in a conversation. The task consists of inferring what a person is implying indirectly. In each item, a correct answer gives two points (for a total of 20 points). An additional hint is given in case of an incorrect answer, after which a correct answer gives one point. The HT assesses second-order ToM because it evaluates the ability to interpret indirect communication, which requires understanding the speaker’s belief about how their hint will be perceived by the listener. The task has good validity and has proven sensitive to ToM difficulties in many studies (Corcoran & Frith, 2003).

The Intelligence Quotient (IQ) of all participants was estimated using the Block Design and Vocabulary or Information WAIS-III subtests (Wechsler, 1997).

Family history was assessed with the Family Interview for Genetic Studies (FIGS) (Díaz de Villalvilla et al., 2008; Nurnberger et al., 1994). Following the broad SZ spectrum criteria (Kendler et al., 1995), families were classified by history with binary coding, as having a positive family history when patients had at least one first-or second-degree relative with SZ, affective or non-affective psychosis, or schizotypal or paranoid personality disorder. Thus, family history values are shared by all family members.

NSS were assessed using structured clinical examination, the Neurological Evaluation Scale (NES) (Buchanan & Heinrichs, 1989, Spanish translation of Gurpegui M у L. Pérez Costillas L, 1994). It consists of 26 items and is divided into four subscales: sensory integration, motor coordination, sequencing of complex motor acts, and "others". Sensory integration includes audiovisual integration, bilateral extinction, graphesthesia, right-left confusion, and stereognosis. The subscale motor coordination encompasses dysdiadochokinesis (rapid alternating movements), finger-to-thumb opposition, finger-to-nose test, and tandem walk. The subscale sequencing of complex motor acts comprises the fist-edge-palm, fist-ring, Ozeretski, and rhythm tapping tests. The subscale "others" includes the assessment of hemispheric dominance, eye movement deficits, frontal release signs, and short-term memory. Each item was scored on a three-point scale, i.e., 0: No abnormality, 1 for mild but definitive impairment, and 2 for marked impairment (except for snout and suck reflex, which were scored 0 or 2). The NES shows high interrater reliability, high intraclass correlations for the total score and subscales and good internal consistency for patients with SZ and healthy controls (Mohr et al., 1996)

PA was evaluated using the Premorbid Adjustment Scale (PAS) (Cannon-Spoor et al., 1982), a retrospective interview that focuses on an individual’s social and academic accomplishments before the onset of illness. The PAS evaluates PA during childhood (up to 11 years), early adolescence (12–15 years), late adolescence (16–18 years), and adulthood (19 years and above). It assesses five domains, including sociability and withdrawal, peer relationships, academic performance, adaptation to school, and social-sexual functioning, and rates them from 0 (normal adjustment) to 6 (severe impairment). The study did not include adult PAS data due to concerns about validity (Horton et al., 2015) and the potential bias introduced by measuring after the disease onset. As a result, maladjustment ratings were only calculated for childhood, early adolescence, and late adolescence. Also, according to previous research (Bechi et al., 2020; Bucci et al., 2016), we computed distinct scores for Social and Academic PA at each stage of development. This was achieved by averaging the sociability and withdrawal, peer relationships, and social-sexual functioning items to represent the social domain and educational performance and adaptation to school items to represent the Academic domain. PA data were available for patients and relatives but not for controls.

### 2.3 Statistical analyses

All data were processed using IBM SPSS Statistics Version 29.0 (SPSS IBM, Armonk, NY: IBM Corp).

Sociodemographic and clinical data were compared between groups using ANOVA or chi-squared tests when appropriate.

The effect of age, years spent in education, and IQ on HT and NES performance for each group was tested using the Pearson correlation test. Within groups, differences in HT performance according to sex or family history were tested using Student’s t-test. From all these analyses, sex and family history showed a significant effect in relatives on HT (p = 0.046 and p = 0.002, respectively), and age and IQ on NES (p=0.029 and p=0.003) in relatives. According to these analyses age, sex and IQ were added as covariates in all the subsequent analyses, and family history in the patients’ and siblings’ intragroup analyses (not in controls as in this group it is a constant=0).

HT performance, NSS and PAS differences between patients and their siblings were assessed through linear mixed models (LMMs) with family member (patients/sibling) as the fixed-effect factor, sex, age and IQ as fixed-effect covariates, and family as the random effect (subjects nested within families). When the analyses included non-related groups (patients vs. controls and siblings vs. controls), the same models were used for the comparison without including the family random effect.

The impact of the markers of altered neurodevelopment (NSS and PA) on HT performance, when taking into account the group and the family, was assessed using a linear mixed model, with HT as the dependent variable, family status (patients/sibling) as the fixed-effect factor, sex, age, IQ and neurodevelopment variables as fixed-effect covariates and family as the random effect (when the analyses included related groups (patients vs. siblings).

Within patients, siblings, and controls, the relationship between HT performance and neurodevelopmental markers was tested using linear regressions (adjusted for age, sex, IQ, and family history).

## 3. RESULTS

### 3.1 Sample characteristics

Their DSM-IV-TR diagnoses were SZ (n=32) and psychotic disorder not otherwise specified (n=6). Patients’ mean age at onset was 22.12 (SD=3.83), and the mean duration of illness was 26.40 months (SD=24.44). All were treated with antipsychotic monotherapy: 94.7%, with second-generation antipsychotic (24 risperidone, five olanzapine, three amisulpride, two ziprasidone, two clozapine) and 5.3% with haloperidol.

The sample’s sociodemographic and clinical features are presented in Table 1. Patients had a larger percentage of men and were younger than siblings and controls. Also, patients showed a lower IQ and fewer years in education compared to controls but did not show differences with siblings.

**Table 1.** Sample description and statistical comparisons between patients (P), siblings (S) and controls (C). Proportion (%) or mean scores (standard deviation) are given. Differences between groups were tested with ANOVA.

|  | **Patients**<br>**(n=38)** | **Siblings**<br>**(n=26)** | **Controls**<br>**(n=47)** | **ANOVA F/$\chi^2$**<br>**(p)** | **Post-hoc**<br>**significant**<br>**differences** |
| --- | --- | --- | --- | --- | --- |
| **N male/female** | 28/10 | 11/15 | 25/22 | 6.89 (0.032) | P>S |
| **Age at interview** | 24.92 (3.90) | 28.79 (5.22) | 27.46 (4.51) | 6.38 (0.002) | S>P, C>P |
| **Years of education** | 13.43 (3.12) | 14.81 (4.14) | 16.30 (3.24) | 7.19 (0.001) | C>P |
| **Estimated**<br>**Intelligence**<br>**Quotient (IQ)** | 90.03<br>(15.23) | 94.15<br>(15.04) | 107.15 (11.00) | 18.35<br>(<0.001) | C >S, C>P |

### 3.2 Neurological Soft Signs in SSD patients relative to their siblings and healthy individuals

Differences in NSS Motor Coordination were found between patients and siblings and between patients and controls (F=5.17 p=0.027 and F=9.38 p= 0.003, respectively) (Table 2, Figure 1). Patients presented higher scores than relatives (estimated mean difference = 0.96) and controls (estimated mean difference = 1.11). Although the mean score of siblings was higher than that of controls, the difference was not statistically significant (see supplementary material for linear mixed model complete results of patients/siblings, patients/controls, siblings/controls; Tables S1, S2, S3 respectively).

**Table 2.** ToM and NSS: clinical description and statistical comparisons between patients (P), siblings (S) and controls (C). Mean scores (standard deviation) are given. Linear Mixed Models (LMM) for pairwise group comparisons with sex, age and IQ as fixed-effect covariates and subjects nested within families when related groups were analysed. Only significant differences are given.

|  | **Patients**<br>**(n=38)** | **Siblings**<br>**(n=26)** | **Controls**<br>**(n=47)** | **Group**<br>**Comparison**<br>**(LMM)** |
| --- | --- | --- | --- | --- |
| **HT** | 15.74(4.07) | 18.46 (1.42) | 18.38 (1.68) | P<S*, P<C |
| **NSS Sensory**<br>**Integration** | 1.71 (1.66) | 1.62 (1.33) | 1.40 (1.42) |  |
| **NSS Motor**<br>**Coordination** | 1.34 (1.81) | 0.42 (0.81) | 0.18 (0.44) | P>S*, P>C** |
| **NSS Sequencing**<br>**complex motor**<br>**acts** | 3.68 (2.70) | 1.69 (1.83) | 0.62 (1.15) | P>S***, S>C*, P>C*** |

\* p< 0.05, \*\* p< 0.01, \*\*\* p<0.001

The comparison of NSS complex motor sequencing between patients and siblings and between patients and controls showed significant group effects (F=15.04, p < 0.001 and F=23.60, p < 0.001, respectively). Patients presented higher scores than relatives (estimated mean difference=2.23) and controls (estimated mean difference=2.72). Also, siblings presented higher scores than controls (F=4.99, p=0.029, estimated mean difference = 0.90) (Table 2) (see supplementary material for the complete results of the linear mixed model of patients/siblings, patients/controls, and siblings/controls Tables S4, S5, and S6, respectively).

The NSS sensory integration scores of patients, siblings and controls did not differ significantly (see supplementary Tables S7-S9).

### 3.3 Premorbid adjustment in SSD patients relative to their siblings

Patients and siblings showed significant differences in social PA during childhood and late adolescence (F=4.83, p=0.032 and F=13.03, p<0.001, respectively), with patients scoring higher than siblings (adjusted mean difference 0.68 and 0.86, respectively). Also, patients showed worse total PA in late adolescence (estimated mean difference = 3.81, F = 10.81, p=0.002) (Table 3) (see supplementary Tables S10-S17).

**Table 3.** Premorbid adjustment Scale (PAS) in patients (P) and siblings (S): clinical description and statistical comparisons between groups. Mean scores (standard deviation) are given. Linear Mixed Model (LMM) for group comparisons with sex, age and IQ as fixed-effect covariates and subjects nested within families. Only significant differences (p<0.05) comparisons are given.

|  |  | **Patients**<br>**(n=38)** | **Siblings**<br>**(n=23)** | **Group**<br>**comparison**<br>**(LMM)** |
| --- | --- | --- | --- | --- |
| **Childhood PAS<sup>a</sup>** | **Total** | 5.05 (3.71) | 3.13 (2.49) |  |
|  | **Social** | 1.05 (1.16) | 0.39 (0.62) | P>S * |
|  | **Academic** | 1.47 (1.09) | 1.17 (0.81) |  |
| **Early adolescence PAS<sup>a</sup>** | **Total** | 7.86 (4.16) | 4.74 (3.19) |  |
|  | **Social** | 1.22 (1.01) | 0.68 (0.67) |  |
|  | **Academic** | 2.01 (1.21) | 1.35 (1.00) |  |
| **Late adolescence PAS<sup>a</sup>** | **Total** | 10.24 (4.72) | 5.00 (2.80) | P>S ** |
|  | **Social** | 1.66 (0.96) | 0.65 (0.56) | P>S *** |
|  | **Academic** | 2.64 (1.73) | 1.52 (1.14) |  |

\* $p < 0.05$ , \*\* $p < 0.01$ , \*\*\* $p < 0.001$

### 3.4 Analysis of ToM performance in patients with SSD relative to their first-degree relatives and healthy individuals and its association with neurodevelopmental markers

HT scores are given in Table 2. A comparison of HT performance between patients and siblings and between patients and controls showed significant group effects (F=5.67, p=0.02) and (F=3.95, p=0.05), respectively. Patients presented lower scores than relatives (estimated mean difference = 2.26) and controls (estimated mean difference = 1.68). The scores of the siblings and controls did not differ significantly (see supplementary Tables S20-S22).

Next, we included the neurodevelopmental markers to analyse their effect on the HT performance differences between SSD patients and siblings. No significant effects of NSS or PA were detected and HT performance differences between the groups remained significant;, however, in most models, the difference in HT means between the groups decreased as compared to the initial model (see supplementary Tables S23-S24).

When analysing the association between HT performance and neurodevelopmental markers within each group, no relationship emerged between HT and any variable of PA or NSS (adjusted by age, sex and IQ). Similarly, no such a relation was observed when adding family history to the patients’ and siblings’ models.

## 4 DISCUSSION

This study aimed to enhance the understanding of ToM variability by examining whether it is affected by early indicators of neurodevelopmental deviances, such as NSS and PA.

Our previous research (Giralt-López et al., 2020, 2024) has shown that, in addition to the more severe ToM deficits observed in patients compared to their relatives and controls, ToM performance may also be influenced among healthy individuals by clinical vulnerability factors, such as schizotypy, basic symptoms, and psychotic-like experiences. This suggests that ToM deficits are not only associated with SZ but also extend to its clinical vulnerability, indicating that ToM could serve as an endophenotypic marker and, therefore, could help identify particularly susceptible individuals, such as those with a genetic predisposition (e.g., relatives). In this sense, family-based studies have been considered a suitable design to deepen the potential role of ToM performance as a vulnerability marker. However, the inconclusive results obtained until now could be attributed to the fact that most of them have focused on first-degree relatives, with limited research specifically targeting siblings of individuals with SZ. Then, the current study adopts a sib-pair approach to control for cohort-related effects relevant to ToM competence and focuses on extending the analysis of ToM variability by assessing the effect of neurodevelopmental markers, given the widespread acceptance of the neurodevelopmental model in SZ (Birnbaum & Weinberger, 2017; Jones & Murray, 1991; Rapoport et al., 2012). Accordingly, NSS and PA were selected to represent the neurobiological basis of the disease and functional development before SZ onset.

First, our approach shows stronger evidence of neurodevelopmental deviations in patients compared to siblings and controls, particularly in specific domains of NSS, such as motor coordination and sequencing of complex motor acts. More concretely, concerning the latter, siblings exhibited an intermediate position between patients and controls, which would support its potential role as the best endophenotypic marker among the three analysed dimensions. In this sense, the specificity of motor domain deviations in NSS could be understood by recognising that motor deficits were linked to abnormalities in brain regions commonly implicated in schizophrenia, such as the cerebellum, basal ganglia, temporal lobe or prefrontal cortex (Luvsannyam et al., 2022). However, research on this field could not report a brain structural specificity for any NSS subscale (Rathod et al., 2020b; Zhao et al., 2014).

These results are aligned with meta-analytic data revealing significant differences in NSS prevalence among SZ patients, their non-psychotic relatives, and healthy controls, supporting the idea that NSS are familial and associated with the illness (Chan, Xu, Heinrichs, Yu, & Gong, 2010). More recent evidence consistently shows that NSS are more prevalent in patients with SZ than in their first-degree relatives, and they are more prevalent in first-degree relatives than in healthy controls, including findings in neuroleptic-naive first-episode patients, further validating NSS as valuable endophenotypes (reviewed by Petrescu et al., 2023; Tsapakis et al., 2023).

More concretely, focusing on specific dimensions, previous studies have shown that the motor coordination subscale is significantly more affected in patients compared to controls, regardless of the illness duration (Chan, Xu, Heinrichs, Yu, & Wang, 2010; Nathani et al., 2023). In studies that included first-degree relatives, findings indicate that patients score higher in motor coordination and complex motor tasks than relatives and controls. However, most of these studies have focused on older patients with a more chronic form of the illness (Chan, Xu, Heinrichs, Yu, & Gong, 2010; Petrescu et al., 2023). Two studies used samples similar to ours, with young patients in early disease stages. One study found increased NSS in motor coordination and sequencing in patients compared to relatives and controls, but no significant intermediate position for siblings (Cuesta et al., 2018). The second study did show an intermediate position for siblings but used a different NSS scale, hampering direct comparisons (Kong et al., 2022).

The fact that NSS are more present in relatives and, to a lesser extent, in controls dissuades the idea that they could be caused by antipsychotic treatments. Furthermore, most studies have not found associations between antipsychotic dosage and the severity of NSS (Compton et al., 2015; Hirjak et al., 2015; Peralta et al., 2010) although NSS tend to decrease in patients who respond adequately to treatment (Bachmann et al., 2014).

Focusing on the second indicator of altered neurodevelopment, PA, our findings, consistent with previous research, show poorer PA in patients than healthy siblings (Shapiro et al., 2009). This could have significant implications for monitoring individuals at high genetic risk, such as the children of those with SSD. Early identification of these high-risk subgroups could facilitate better management of environmental risk factors and enable closer monitoring for early signs of psychosis, potentially reducing the duration of untreated psychosis, which is associated with poorer outcomes.

After group differences analyses, our goal was to demonstrate whether the variability in these markers – indicative of early neurodevelopmental deviations -accounts for differences in ToM skills, a cognitive variable also linked to neurodevelopment (Baron-Cohen et al., 1999; Damasceno, 2020). When comparing HT performance between SSD patients and siblings, adjusted for NSS or PA, our study could not demonstrate any significant role of these neurodevelopmental markers contributing to ToM variability across groups. Additionally, beyond the between-groups effect, we also tested the association of these two neurodevelopmental markers with ToM within each group. However, unlike previous studies that identified a correlation between NSS and poorer ToM performance in patients (Herold et al., 2019; Romeo et al., 2014), our research does not find such relationship in the within-group analyses of patient, sibling, and controls. Then, our data does not support that specific NSS and ToM deficits might share common neural mechanisms as proposed by these previous studies. Although in all studies age of onset and years of education are similar, our sample is characterised by including patients in the early stages of the disease (less than 3 years of evolution), while previous studies (Herold et al., 2019; Romeo et al., 2014) included older patients with chronic SZ of more than 10 years of evolution. It is also interesting to note that both studies adjusted the analyses by age and years of education but not by IQ, unlike our study, which included IQ in the analyses considering the reported association of neurocognitive symptoms (including attention, information processing, processing speed, reasoning and problem-solving, social cognition, working memory, and verbal and visual learning and memory) with NSS, reviewed in (Tsapakis et al., 2023). Accordingly, our findings are consistent with those reported in other developmental disorders, such as ADHD, that although they could confirm NSS as markers of atypical neurodevelopment and predictors of the severity of functional impairment, did not find a correlation with ToM abilities (Pitzianti et al., 2017).

Similarly to what was observed in the case of the NSS, our study could not add evidence supporting the recently reviewed and described relationship between ToM and PA in patients with schizophrenia (Punsoda-Puche et al., 2024). To the best of our knowledge, only one family-based study reports altered academic PA in relatives, including both parents and siblings. Although this study evaluated social cognition in the patient group (using a composite score that was not specific to ToM skills or their relationship with PA) it did not explore this dimension within the relatives’ group (Bucci et al., 2018).

In summary, this study did not find evidence of a correlation between ToM and markers of altered neurodevelopment across the patient, sibling, and control groups, contrary to expectations. This is despite the well-documented ontogenic development of ToM abilities in the early stages of development (Baron-Cohen et al., 1985; Damasceno, 2020). Then, these results suggest they may potentially rely on different neurodevelopmental-related pathways. In this sense, it is remarkable that ToM, NSS, and PA all represent indirect approaches to understanding neurodevelopment, but from complementary perspectives. Specifically, NSS likely serve as a marker closely related to the central nervous system substrate, while PA acts as an indirect functional marker of altered neurodevelopment, and ToM reflects the cognitive level.

Attempts to describe the neurobiological roots of ToM should consider additional markers of altered neurodevelopment that correlate with ToM deficits. This could help identify a potential shared neurobiological substrate. With special focus on the relatives’ group, who have a greater genetical risk (Gottesman, 1991), a comprehensive approach to define subgroups of family members at higher risk could involve integrating neurodevelopmental markers with indicators of clinical vulnerability, such as levels of schizotypy. Prior evidence has shown a relationship between these two types of risk markers, more concretely NSS (especially motor signs), were associated with some schizotypal dimensions in siblings of patients with schizophrenia (Mechri et al., 2010). Furthermore, it would be valuable to explore the role of environmental factors that may interact with early neurodevelopmental alterations and mediate reduced ToM abilities. Considering these interactions may provide a more comprehensive understanding of the mechanisms influencing ToM deficits.

Finally, it would be interesting if future studies could longitudinally examine the relationship between ToM and NSS. It would be of particular interest to evaluate this relationship also in patients in the acute phase, considering the potential attenuating effect that the remission of acute symptoms can have (at the same time related to the initiation of antipsychotic treatment) (Bachmann et al., 2014).

To properly understand our findings, it is essential to acknowledge the study’s strengths and limitations. Strengths include using a family-based design with an affected-unaffected sibling approach, a control group for most variables, the use of the most suitable tool for evaluating Theory of Mind (ToM) within the psychosis continuum (Ludwig et al., 2017), and the inclusion of early neurodevelopmental markers. These aspects of our study help address the ongoing discussion about whether ToM characteristics are state or trait-related and provide insights into the complex traits linked to SZ risk. However, despite having a sample size comparable to previous studies, a significant limitation is that our sample size is still relatively small, which is a common challenge in family-based research that requires large, well-characterised families. Additionally, while ToM is a crucial element of social cognition, our study did not examine other areas, such as emotion processing and social knowledge and treatment.

In conclusion, in our sample, while patients showed more evidence of neurodevelopmental deviances (assessed through NSS and PA) than siblings and controls, such differences do not explain ToM variability. These findings encourage further exploration of endophenotypic markers or combinations of markers that can enhance the stratification of risk for conversion to psychosis, particularly within the general population and among siblings or children of patients who face an elevated risk due to their genetic predisposition. A more precise definition of these high-risk groups would enable the development of targeted interventions to mitigate this risk or concentrate efforts on early detection, ultimately improving the already proven cost-effectiveness of early psychosis care programs (Ologundudu et al., 2023).

## 6 AUTHORS’ CONTRIBUTIONS

MG-L: Conceptualization, Data curation, Formal analysis, Methodology, Visualisation, Writing – original draft. SM: Conceptualization, Investigation, Methodology, Resources, Writing – review & editing. SC: Investigation, Writing – review & editing. MM: Investigation, Writing – review & editing. AS-M: Investigation, Writing – review & editing. NH: Investigation, Writing – review & editing, MOK: Conceptualization, Funding acquisition, Writing – review & editing. LF: Conceptualisation, Funding acquisition, Resources, Writing – review & editing. MF-V: Conceptualisation, Data curation, Formal analysis, Funding acquisition, Investigation, Methodology, Resources, Supervision, Writing – original draft

## ACKNOWLEDGMENTS

We are deeply grateful to all participants whose generosity made this work possible.

## 8 DATA AVAILABILITY STATEMENT

The dataset generated for this study is available on request to the corresponding authors.

## 9 ETHICS STATEMENT

The study involving humans was approved by the Ethics Committee of the University of Barcelona (IRB0003099). The study was conducted in accordance with the local legislation and institutional requirements. The participants provided their written informed consent to participate in the study.

