## Supplementary Material for "Theory of Mind Variability in Schizophrenia: A Neurodevelopmental Perspective through Neurological Soft Signs and Premorbid Adjustment"

**SUPPLEMENTARY MATERIAL PAPER 3**

**Table S1.** Results of the linear mixed model, with NSS motor coordination as the dependent variable, family status (patients / sibling) as the fixed-effect factor, sex, age and IQ as fixed-effect covariates and family as the random effect (subjects nested within families).

|  | **Estimated effect** | **F** | **p-value** |
| --- | --- | --- | --- |
| **Age at interview** | -.032189 | .535 | .467 |
| **Sex** | .440289 | 1.116 | .295 |
| **IQ** | -.002087 | .026 | .871 |
| **status** | .962734 | 5.167 | .027 |

**Table S2.** Results of the linear mixed model, with NSS motor coordination as the dependent variable, family status (patients / controls) as the fixed-effect factor and sex, age and IQ as fixed-effect covariates.

|  | **Estimated effect** | **F** | **p-value** |
| --- | --- | --- | --- |
| **Age at interview** | .809734 | .600 | .441 |
| **Sex** | -.026924 | .437 | .511 |
| **IQ** | .207623 | .028 | .868 |
| **status** | -.001911 | 9.379 | .003 |

**Table S3.** Results of the linear mixed model, with total NSS motor coordination as the dependent variable, family status (siblings / controls) as the fixed-effect factor and sex, age and IQ as fixed-effect covariates.

|  | **Estimated effect** | **F** | **p-value** |
| --- | --- | --- | --- |
| **Age at interview** | .027956 | 3.092 | .083 |
| **Sex** | -.047871 | .096 | .758 |
| **IQ** | -.003244 | .277 | .601 |
| **status** | .161116 | .875 | .353 |

**Table S4.** Results of the linear mixed model, with NSS complex motor sequencing as the dependent variable, family status (patients / sibling) as the fixed-effect factor, sex, age and IQ as fixed-effect covariates and family as the random effect (subjects nested within families).

|  | **Estimated effect** | **F** | **p-value** |
| --- | --- | --- | --- |
| **Age at interview** | -.073227 | 1.423 | .238 |
| **Sex** | 1.618753 | 7.454 | .008 |
| **IQ** | -.042235 | 5.798 | .019 |
| **status** | 2.224973 | 15.040 | <.001 |

**Table S5.** Results of the linear mixed model, with NSS complex motor sequencing as the dependent variable, family status (patients / controls) as the fixed-effect factor and sex, age and IQ as fixed-effect covariates.

|  | **Estimated effect** | **F** | **p-value** |
| --- | --- | --- | --- |
| **Age at interview** | -.069654 | 1.809 | .182 |
| **Sex** | .521773 | 1.190 | .279 |
| **IQ** | -.016452 | .874 | .353 |
| **status** | 2.714575 | 23.606 | <.001 |

**Table S6.** Results of the linear mixed model, with NSS complex motor sequencing as the dependent variable, family status (siblings / controls) as the fixed-effect factor and sex, age and IQ as fixed-effect covariates.

|  | **Estimated effect** | **F** | **p-value** |
| --- | --- | --- | --- |
| **Age at interview** | -.002686 | .005 | .942 |
| **Sex** | -.292850 | .640 | .426 |
| **IQ** | -.015823 | 1.133 | .291 |
| **status** | .899517 | 4.914 | .030 |

**Table S7.** Results of the linear mixed model, with NSS sensory integration as the dependent variable, family status (patients / sibling) as the fixed-effect factor, sex, age and IQ as fixed-effect covariates and family as the random effect (subjects nested within families).

|  | **Estimated effect** | **F** | **p-value** |
| --- | --- | --- | --- |
| **Age at interview** | .054312 | 1.527 | .221 |
| **Sex** | -.072919 | .031 | .862 |
| **IQ** | -.033079 | 6.685 | .012 |
| **status** | .161136 | .146 | .704 |

**Table S8.** Results of the linear mixed model, with NSS sensory integration as the dependent variable, family status (patients / controls) as the fixed-effect factor and sex, age and IQ as fixed-effect covariates.

|  | **Estimated effect** | **F** | **p-value** |
| --- | --- | --- | --- |
| **Age at interview** | .068142 | 3.015 | .086 |
| **Sex** | .024310 | .004 | .947 |
| **IQ** | -.031786 | 5.684 | .019 |
| **status** | -.059033 | .019 | .889 |

**Table S9.** Results of the linear mixed model, with NSS sensory integration as the dependent variable, family status (siblings / controls) as the fixed-effect factor and sex, age and IQ as fixed-effect covariates.

|  | **Estimated effect** | **F** | **p-value** |
| --- | --- | --- | --- |
| **Age at interview** | 7.677899E-5 | .000 | .998 |
| **Sex** | -.320680 | .955 | .332 |
| **IQ** | -.039534 | 9.940 | .003 |
| **status** | -.369839 | .929 | .339 |

**Table S10.** Results of the linear mixed model, with Total PAS during childhood as the dependent variable, family status (patients / sibling) as the fixed-effect factor, sex, age and IQ as fixed-effect covariates and family as the random effect (subjects nested within families).

|  | **Estimated effect** | **F** | **p-value** |
| --- | --- | --- | --- |
| **Age at interview** | .076214 | .678 | .414 |
| **Sex** | -2.534965 | 7.994 | .007 |
| **IQ** | -.058453 | 4.671 | .036 |
| **status** | .989543 | 1.153 | .288 |

**Table S11.** Results of the linear mixed model, with Social PAS during childhood as the dependent variable, family status (patients / sibling) as the fixed-effect factor, sex, age and IQ as fixed-effect covariates and family as the random effect (subjects nested within families).

|  | **Estimated effect** | **F** | **p-value** |
| --- | --- | --- | --- |
| **Age at interview** | .018277 | .369 | .546 |
| **Sex** | -.197511 | .479 | .492 |
| **IQ** | .002145 | .054 | .818 |
| **status** | .675996 | 4.817 | .032 |

**Table S12.** Results of the linear mixed model, with Academic PAS during childhood as the dependent variable, family status (patients / sibling) as the fixed-effect factor, sex, age and IQ as fixed-effect covariates and family as the random effect (subjects nested within families).

|  | **Estimated effect** | **F** | **p-value** |
| --- | --- | --- | --- |
| **Age at interview** | .019115 | .679 | .414 |
| **Sex** | -1.034233 | 20.806 | <.001 |
| **IQ** | -.029939 | 20.556 | <.001 |
| **status** | -.119391 | .277 | .601 |

**Table S13.** Results of the linear mixed model, with Total PAS during early adolescence as the dependent variable, family status (patients / sibling) as the fixed-effect factor, sex, age and IQ as fixed-effect covariates and family as the random effect (subjects nested within families).

|  | **Estimated effect** | **F** | **p-value** |
| --- | --- | --- | --- |
| **Age at interview** | .001278 | .000 | .991 |
| **Sex** | -3.111004 | 8.052 | .007 |
| **IQ** | -.042742 | 1.745 | .193 |
| **status** | 1.607487 | 2.040 | .159 |

**Table S14.** Results of the linear mixed model, with Social PAS during early adolescence as the dependent variable, family status (patients / sibling) as the fixed-effect factor, sex, age and IQ as fixed-effect covariates and family as the random effect (subjects nested within families).

|  | **Estimated effect** | **F** | **p-value** |
| --- | --- | --- | --- |
| **Age at interview** | .008796 | .106 | .746 |
| **Sex** | -.367385 | 1.900 | .174 |
| **IQ** | .008978 | 1.362 | .251 |
| **status** | .400900 | 2.307 | .137 |

**Table S15.** Results of the linear mixed model, with Academic PAS during early adolescence as the dependent variable, family status (patients / sibling) as the fixed-effect factor, sex, age and IQ as fixed-effect covariates and family as the random effect (subjects nested within families).

|  | **Estimated effect** | **F** | **p-value** |
| --- | --- | --- | --- |
| **Age at interview** | -.018300 | .450 | .505 |
| **Sex** | -1.143616 | 18.633 | <.001 |
| **IQ** | -.035953 | 19.698 | <.001 |
| **status** | .018809 | .005 | .945 |

**Table S16.** Results of the linear mixed model, with Total PAS during late adolescence as the dependent variable, family status (patients / sibling) as the fixed-effect factor, sex, age and IQ as fixed-effect covariates and family as the random effect (subjects nested within families).

|  | **Estimated effect** | **F** | **p-value** |
| --- | --- | --- | --- |
| **Age at interview** | .097766 | .699 | .407 |
| **Sex** | -3.512888 | 9.398 | .004 |
| **IQ** | -.039293 | 1.337 | .255 |
| **status** | 3.809762 | 10.812 | .002 |

**Table S17.** Results of the linear mixed model, with Social PAS during late adolescence as the dependent variable, family status (patients / sibling) as the fixed-effect factor, sex, age and IQ as fixed-effect covariates and family as the random effect (subjects nested within families).

|  | **Estimated effect** | **F** | **p-value** |
| --- | --- | --- | --- |
| **Age at interview** | .002181 | .008 | .929 |
| **Sex** | -.315275 | 1.690 | .199 |
| **IQ** | .011709 | 2.938 | .095 |
| **status** | .858495 | 13.303 | <.001 |

**Table S18.** Results of the linear mixed model, with Academic PAS during late adolescence as the dependent variable, family status (patients / sibling) as the fixed-effect factor, sex, age and IQ as fixed-effect covariates and family as the random effect (subjects nested within families).

|  | **Estimated effect** | **F** | **p-value** |
| --- | --- | --- | --- |
| **Age at interview** | .045217 | 1.223 | .274 |
| **Sex** | -1.402569 | 12.770 | <.001 |
| **IQ** | -.036486 | 8.619 | .005 |
| **status** | .620382 | 2.233 | .141 |

**Table S20.** Results of the linear mixed model, with HT as the dependent variable, family status (patients / sibling) as the fixed-effect factor, sex, age and IQ as fixed-effect covariates and family as the random effect (subjects nested within families).

|  | **Estimated effect** | **F** | **p-value** |
| --- | --- | --- | --- |
| **Age at interview** | .045483 | .223 | .638 |
| **Sex** | .441776 | .243 | .624 |
| **IQ** | .037545 | 1.729 | .194 |
| **status** | -2.254973 | 5.673 | .020 |

**Table S21** Results of the linear mixed model, with HT as the dependent variable, family status (patients / controls) as the fixed-effect factor and sex, age and IQ as fixed-effect covariates.

|  | **Estimated effect** | **F** | **p-value** |
| --- | --- | --- | --- |
| **Age at interview** | .004594 | .003 | .953 |
| **Sex** | .224831 | .097 | .757 |
| **IQ** | .053156 | 3.992 | .049 |
| **status** | -1.678160 | 3.945 | .050 |

**Table S22** Results of the linear mixed model, with HT as the dependent variable, family status (siblings / controls) as the fixed-effect factor and sex, age and IQ as fixed-effect covariates.

|  | **Estimated effect** | **F** | **p-value** |
| --- | --- | --- | --- |
| **Age at interview** | -.042582 | 1.164 | .285 |
| **Sex** | .384096 | .954 | .333 |
| **IQ** | .015977 | 1.164 | .288 |
| **status** | .209032 | .202 | .655 |

**Table S23.** Results of the linear mixed model, with HT as the dependent variable, family status (patients / sibling) as the fixed-effect factor, sex, age, IQ and neurodevelopment variables as fixed-effect covariates and family as the random effect (subjects nested within families).

| **Neurodevelopment variable**  **(ND variable)** | **F of the ND variable** | **p of the ND variable** | **Estimated Mean difference** | **F of the model** | **p of the model** |
| --- | --- | --- | --- | --- | --- |
| **NSS Sensory Integration** | 0.855 | 0.359 | -2.292 | 5.840 | 0.019 |
| **NSS Motor Coordination** | 1.407 | 0.240 | -1.940 | 3.914 | 0.053 |
| **NSS Sequencing complex motor acts** | 0.014 | 0.907 | -2.304 | 4.880 | 0.031 |
| **Academic PAS Childhood** | 0.189 | 0.665 | -2.083 | 4.125 | 0.047 |
| **Academic PAS Early Adolescence** | 1.070 | 0.306 | -2.211 | 4.739 | 0.034 |
| **Academic PAS Late Adolescence** | 0.000 | 0.996 | -2.202 | 4.430 | 0.040 |
| **Social PAS Childhood** | 0.885 | 0.351 | -1.780 | 2.813 | 0.099 |
| **Social PAS Early Adolescence** | 1.070 | 0.306 | -1.946 | 3.464 | 0.068 |
| **Social PAS Late Adolescence** | 1.336 | 0.253 | -1.580 | 1.896 | 0.174 |

**Table S24.** Results of the linear mixed model, with HT as the dependent variable, family status (patients / controls) as the fixed-effect factor, sex, age, IQ and neurodevelopment variables as fixed-effect covariates.

| **Neurodevelopment variable** | **F of the ND variable** | **p of the ND variable** | **Estimated Mean difference** | **F of the model** | **P of the model** |
| --- | --- | --- | --- | --- | --- |
| **NSS Sensory Integration** | 0.918 | 0.341 | -1.666 | 3.881 | 0.052 |
| **NSS Motor Coordination** | 2.498 | 0.118 | -1.216 | 1.834 | 0.180 |
| **NSS Sequencing complex motor acts** | 0.193 | 0.662 | -1.881 | 3.787 | 0.055 |
